# Effectiveness of analgesic ear drops as add-on treatment to usual care in children with acute otitis media: the OPTIMA pragmatic randomised controlled trial

**DOI:** 10.1101/2025.09.24.25336532

**Authors:** JLH de Sévaux, RAMJ Damoiseaux, AGM Schilder, PA Zuithoff Nicolaas, RP Venekamp

**Author notes:** **Address correspondence to:** Joline L.H. de Sévaux, Julius Center for Health Sciences and Primary Care, University Medical Center Utrecht, Str. 6.131, P.O. Box 85500, 3508 GA Utrecht, the Netherlands [ ].

## Abstract

**Background:** Clinical practice guidelines recommend oral analgesics for all children with acute otitis media (AOM), and antibiotics for selected cases. Analgesic ear drops could be a promising add-on treatment, but evidence for their effectiveness is limited. Despite being available over-the-counter, they are rarely used in daily practice.

**Aim:** To establish whether analgesic ear drops added to usual care provides superior ear pain relief over usual care alone in children presenting to general practice with AOM.

**Design and setting:** Pragmatic, open label, two arm, individually randomised, superiority trial in 35 general practices in the Netherlands between October 2021 and January 2024.

**Methods:** Children with AOM and ear pain were randomly allocated to either 1) lidocaine hydrochloride 5mg/g ear drops 1-2 drops up to six times daily for a maximum of 7 days in addition to usual care (oral analgesics with or without antibiotics) or 2) usual care. The primary outcome, parent-reported ear pain score (0-10) over the first 3 days, was analysed with a linear regression model accounting for repeated measurements, adjusting for time, baseline ear pain score, age ≤2 vs >2 years, uni-vs bilateral AOM, and baseline antibiotic prescribing. Secondary outcomes were reported descriptively.

**Results:** With 29 of a planned 300 children enrolled between Oct 2021 and Jan 2024 (15 intervention, 14 control; mean age 2.6 years (SD 1.5), 62% boys), the trial was ended prematurely due to slow accrual. Analyses were conducted for 28 participants with complete data. No significant difference in mean parent-reported pain scores over the first three days was observed between children receiving analgesic ear drops in addition to usual care and those receiving usual care alone (mean difference 0.14, 95% CI -2.00 to 2.28). Local discomfort and difficulties during administration of the analgesic ear drops were reported in 71.4% (10/14) and 50% (7/14) of children, respectively.

**Reflections:** Early termination of this trial does not allow for conclusions on the effectiveness of analgesic ear drops as add-on treatment in children with AOM. Our experience underscores the substantial challenges of delivering prospective trials in general practice, and underlines the need for collaborative and viable recruitment infrastructures. Combining individual participant trial data from existing trials may help provide more robust conclusions.

**Trial registration:** The Netherlands Trial Register; NL9500. Date of registration: 28 May 2021.

## INTRODUCTION

Acute otitis media (AOM) is one of the most common childhood diagnoses and a leading cause of antibiotic prescribing in general practice. Ear pain and fever are the main reasons for consultation(1-4), with ear pain persisting for up to eight days.(5, 6) Effective pain relief is therefore key to the management of AOM. Current guidelines recommend oral analgesics for all children with AOM and antibiotics for selected cases.(7, 8) In day to day practice, many children with AOM receive antibiotics.(2, 9) Side effects of antibiotics occur in 10% of children, and widespread use of antibiotics for AOM contributes to antimicrobial resistance.(10, 11) Optimised pain relief may help lower antibiotic prescribing in children with AOM, and reduce exposure to these side effects.

There is some evidence that although paracetamol or ibuprofen reduce ear pain, pain relief is often insufficient with up to 10% of children still experiencing ear pain at 48 hours despite their use.(12) This highlights the need to explore additional treatment options that could safely and effectively alleviate ear pain and reduce reliance on antibiotics in childhood AOM.(13) Analgesic ear drops may offer a low-cost and low-risk addition to usual care.(14) Despite their over-the-counter availability, they are infrequently used in the Netherlands, with parents reporting use in only about 5% of children with AOM.(15) Also, evidence on their effectiveness in AOM is limited and comes from small trials, using different active ingredients, such as benzocaine, phenazone, lignocaine, or lidocaine.(14, 16-18)

We therefore initiated a randomised controlled trial (RCT) to establish if analgesic ear drops added to usual care (oral analgesics with or without antibiotics) provide superior ear pain relief over usual care alone in children presenting to general practice with AOM and ear pain.

## METHODS

### Study design and setting

From October 2021 to January 2024, we conducted a pragmatic, open label, two arm, individually randomised, superiority trial in 35 general practices (114 general practitioners (GPs)) in the region Utrecht, the Netherlands. Due to slow participant accrual, the trial was ended prematurely on 10 January 2024.

The trial’s rationale and details of its design have been reported in detail elsewhere.(19) The trial protocol was approved by Medical Research Ethics Committee NedMec (21-447/G-D) and trial results were reported according to the Consolidated Standards of Reporting Trials (CONSORT) guideline.(20)

### Participants

Children aged 1-6 years presenting to their GP with parent-reported ear pain in the previous 24 hours and diagnosed with AOM were eligible for trial participation. Children with ear wax obscuring visualisation of the tympanic membrane, those with a (suspected) tympanic membrane perforation or ventilation tube present, and those who were very unwell or required hospitalisation were excluded. Detailed inclusion and exclusion criteria are presented in Box 1.

### Study procedures and data collection

#### Recruitment

GPs informed parents of potentially eligible children about the trial, took consent to share their contact details with the study team at the UMC Utrecht and provided a study information leaflet. Upon receipt of the contact details, the trial doctor contacted parents by phone to provide detailed information about the study and scheduled a home visit on the same day for those who provisionally agreed to participate.

#### Study group assignment

An independent data manager generated a computer-generated randomisation sequence prior to trial initiation, stratified for age (<2 versus ≥2 years), AOM laterality (unilateral versus bilateral), and baseline antibiotic prescribing (yes versus no). After written informed consent and completion of all baseline assessments, children were randomly assigned to the intervention or control group via a secure web-based system ensuring concealed allocation. Assignment was balanced in a 1:1 ratio for the two study groups.

Children randomised to the intervention group received lidocaine hydrochloride 5mg/g (Otalgan^®^) ear drops, 1-2 drops per administration, up to six times daily for a maximum of 7 days, in addition to usual care (oral analgesics, with or without antibiotics). Parents were instructed to administer the drops when they felt their child was suffering ear pain, while not exceeding the maximum daily dosage or the maximum treatment duration of 7 days, and to discontinue treatment immediately if otorrhoea developed. Those allocated to the control group received usual care alone. Any treatment decisions other than the use of analgesic ear drops, i.e. antibiotics and oral analgesics use, were left to the GP’s discretion in both groups.

#### Baseline and follow-up data collection

At the home visit, the trial doctor collected demographic and disease-specific data and recorded otoscopic findings. Otoscopy was performed with a Welch Allyn MacroView Otoscope^®^. During the 4 week follow-up, parents kept a diary of AOM-related symptoms including ear pain scores and fever recordings, treatment adherence, prescribed and over-the-counter (OTC) medication use, AOM-related health care resource use, adverse events and complications of AOM.

### Outcomes

The primary outcome was parent-reported ear pain score over the first three days after trial inclusion. To this end, parents recorded their child’s overall ear pain for each day using a 0-10 validated numerical rating scale.(21)

Secondary outcomes included: antibiotic use (yes/no) in the first 7 days, oral analgesic use (yes/no) in the first 7 days, number of days with ear pain during follow up, number of GP reconsultations with or without subsequent antibiotic prescribing during follow-up, overall symptom burden (crying/distress, disturbed sleep, interference with normal activity, appetite, fever and hearing problems) measured on a 0-6 Likert scale in the first 7 days (14, 21), generic quality of life of the child at baseline and 4 weeks assessed using the 47-item short-form of the Infant Toddler Quality of Life Questionnaire (ITQOL-SF47)(22), disease-specific quality of life of the child at baseline and 4 weeks assessed using the Otitis media-6 (OM-6) questionnaire(23), adverse events during follow-up, and complications of AOM during follow-up. For the ITQOL-SF47, we report results for three key domains selected for their relevance to the study context: physical functioning, general health perceptions, and parental emotional impact. For the symptom burden, we calculated a daily total score by summing parent-reported severity scores across six symptoms (range 0–6 per symptom): crying/distress, disturbed sleep, activity interference, appetite, fever, and hearing problems. Higher scores indicated greater severity.

### Patient and public involvement

A panel of eight parents was brought together for this trial. Through online panel meetings they helped shape the grant proposal and study protocol, with particular input on the outcomes, minimally important clinical difference for the primary outcome, recruitment strategy, and data collection procedures.

### Sample size calculation

The study aimed to demonstrate that analgesic ear drops offer superior parent-reported ear pain relief over the first three days after trial inclusion compared to usual care. Based on previous data and input from our parent panel, a 0.87-point reduction in pain (20%) was considered clinically meaningful (14, 15). To detect this with 90% power and a 5% significance level, 126 children per group were needed. Accounting for 20% attrition, we aimed to recruit 300 participants.

### Statistical analysis

All analyses were performed according to the intention-to-treat (ITT) principle. Baseline characteristics were summarized descriptively for each treatment group. The primary outcome was analysed with a covariance pattern model (CPM). A CPM is a linear regression model that includes a residual covariance (i.e. a GEE-type) matrix to account for repeated measurements in the same patients over time. In addition to the treatment arm, the analysis was adjusted for time (i.e. day 1, 2 and 3), the baseline ear pain score, age (≤2 vs >2 years), AOM laterality (uni-vs bilateral), and baseline antibiotic prescribing (yes vs no).(24) In an additional step, potential differences between treatment arms in the reduction of pain scores over time were assessed by including the time by treatment interaction and tested with a likelihood ratio test (LRT). Results were expressed as differences in mean pain scores with 95% confidence intervals (CI). Model assumptions (i.e. distributional assumptions, homoscedasticity) were assessed with residual analyses. Secondary outcomes were reported descriptively. All statistical analyses were performed with IBM SPSS Statistics (version 27.0) and SAS 9.4.

## RESULTS

### Participants

The trial was ended prematurely due to slow accrual, with only 29 of the planned 300 children enrolled. Among the enrolled participants, 15 were randomly assigned to analgesic ear drops in addition to usual care and 14 to usual care alone (Figure 1). Follow-up data were unavailable for one participant in the intervention group; analyses were conducted with 28 participants.

**Figure 1.**
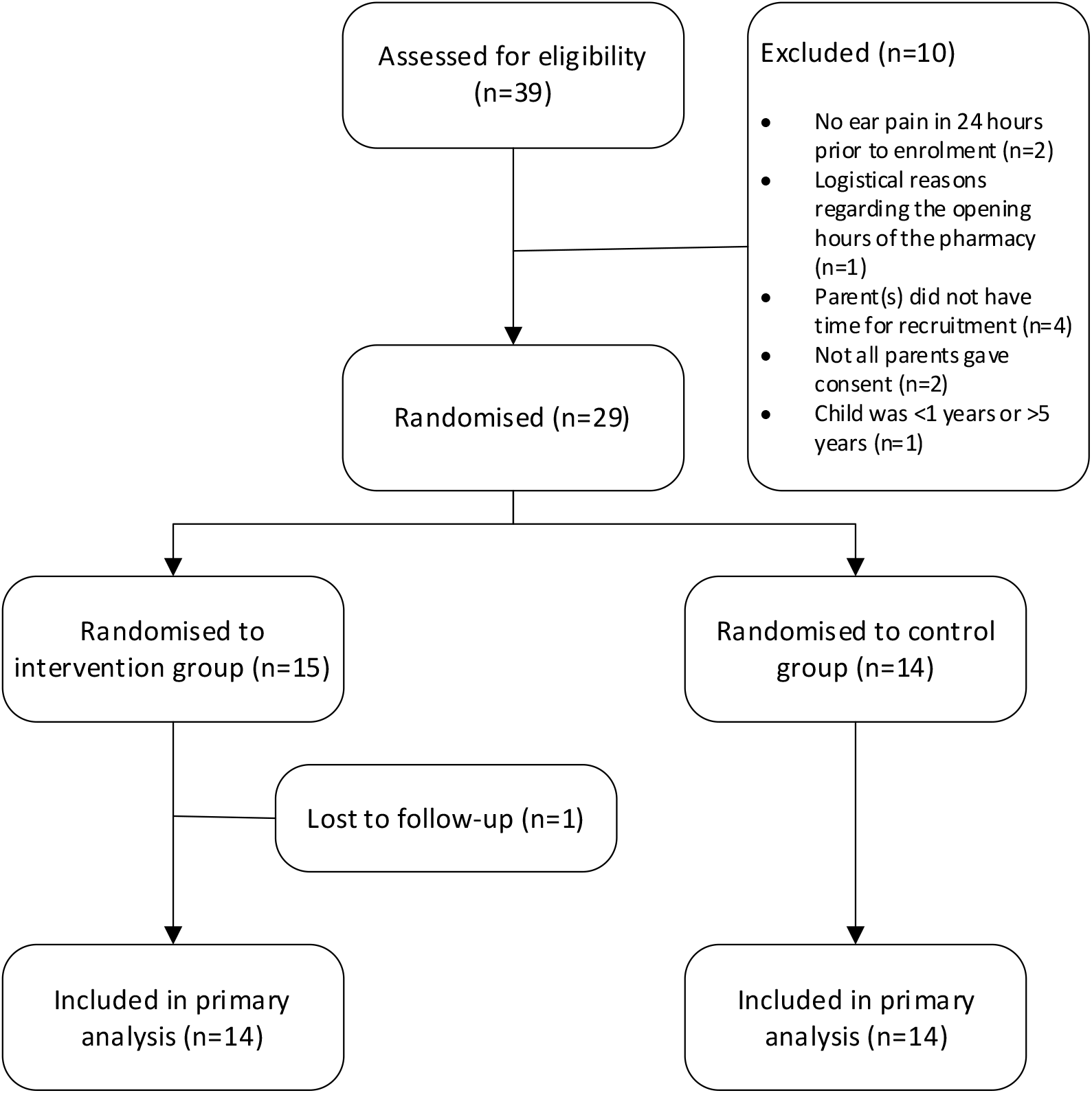
Flowchart

On days with reported ear pain, 73.1% of children in the intervention group received analgesic ear drops, with a mean of 2.21 administrations per day. On days, however, without ear pain, 41.5% of these children received ear drops, with a mean of 0.59 administrations per day. None of the children in the usual care group received analgesic eardrops.

There were some baseline differences between the groups: children in the intervention group were slightly younger (2.3 versus 2.9 years), had a higher mean baseline ear pain score (7.7 versus 6.5), and were prescribed antibiotics more frequently (33.3% versus 21.4%) than those in the control group (Table 1).

**Table 1.** Baseline characteristics.

| Characteristic | Intervention (n=15)<br>*mean ±sd or n (%)* | Control (n=14)<br>*mean ±sd or n (%)* | All children (n=29)<br>*mean ±sd or n (%)* |
| --- | --- | --- | --- |
| <u>Age</u> |  |  |  |
| years | 2.3 ±1.44 | 2.9 ±1.54 | 2.6 ±1.50 |
| <2 years | 7 (46.7%) | 4 (28.6%) | 11 (37.9%) |
| <u>Sex</u> |  |  |  |
| Male | 10 (66.7%) | 8 (57.1%) | 18 (62.1%) |
| Baseline ear pain score (1-10) | 7.7 ±1.63 | 6.5 ±1.91 | 7.1 ±1.84 |
| Number of days in pain | 2.8 ± 1.47 | 2.9 ±1.90 | 2.9 ±1.66 |
| Baseline antibiotic prescription | 5 (33.3%) | 2 (14.3%) | 7 (24.1%) |
| Bilateral AOM | 5 (33.3%) | 4 (28.6%) | 9 (31.0%) |
| Household smoking | 3 (20%) | 0 (0%) | 3 (10.3%) |
| Additional children in household | 9 (60%) | 10 (71.4%) | 19 (65.5%) |
| Breastfed at 3 months | 10 (66.7%) | 10 (71.4%) | 20 (69.0%) |
| Temperature | 37.7 ± 0.99 | 38.2 ±1.07 | 37.9 ±1.04 |
| <u>Symptoms (scale 0-6)</u> |  |  |  |
| Crying | 4.5 ±1.36 | 3.3 ±1.90 | 3.9 ±1.72 |
| Disturbed sleep | 4.3 ±1.16 | 3.6 ±2.50 | 3.9 ±1.93 |
| Reduced activity | 2.5 ±2.10 | 3.0 ±1.36 | 2.8 ±1.77 |
| Reduced appetite | 3.1 ±2.22 | 3.2 ±1.81 | 3.2 ±2.00 |
| Fever | 2.6 ±2.26 | 2.9 ±2.45 | 2.7 ±2.31 |
| Hearing problems | 1.1 ±1.41 | 1.9 ±1.70 | 1.5 ±1.57 |

### Primary outcome

Mean ear pain scores decreased over the first three days in both groups. Descriptively, children in the intervention group had higher mean pain scores at each individual time point than those in the control group (Figure 2, Table 2). In primary analysis, there was no significant difference in mean parent-reported pain scores over the first three days between children receiving analgesic ear drops in addition to usual care and those receiving usual care alone (adjusted mean difference 0.14, 95% CI −2.00 to 2.28) (Table 2). The LRT for the time by treatment interaction showed non-significant results (LR Chi^2^= 0.1989, df=2, p=0.91).

**Table 2.** Primary outcome.

| **Ear pain score (1-10)** | **All children (n=28)**<br>**mean ±sd** | **Intervention (n=14)**<br>**mean ±sd** | **Control (n=14)**<br>**mean ±sd** | **Intervention effect**<br>**Unadjusted mean difference (95% CI)** | **Intervention effect**<br>**Adjusted mean difference (95% CI)\*** |
| --- | --- | --- | --- | --- | --- |
| At baseline | 7.1 ±1.84 | 7.7 ±1.63 | 6.5 ±1.91 | - | - |
| Day 1 | 5.6 ±2.90 | 5.9 ±2.84 | 5.3 ±3.02 | - | - |
| Day 2 | 4.3 ±2.79 | 4.8 ±2.67 | 3.8 ±2.91 | - | - |
| Day 3 | 3.4 ±2.88 | 3.7 ±2.97 | 3.0 ±2.86 | - | - |
| Over the first 3 days | - | - | - | 0.76 (95% CI -1.15 to 2.66) | 0.14 (95% CI -2.00 to 2.28) |

CI = confidence interval; \*adjusted for time (i.e. day 1, 2 and 3), the baseline ear pain score, age ( $\leq 2$ vs $>2$ years), AOM laterality (uni- vs bilateral), and baseline antibiotic prescribing (yes vs no).

**Figure 2.**
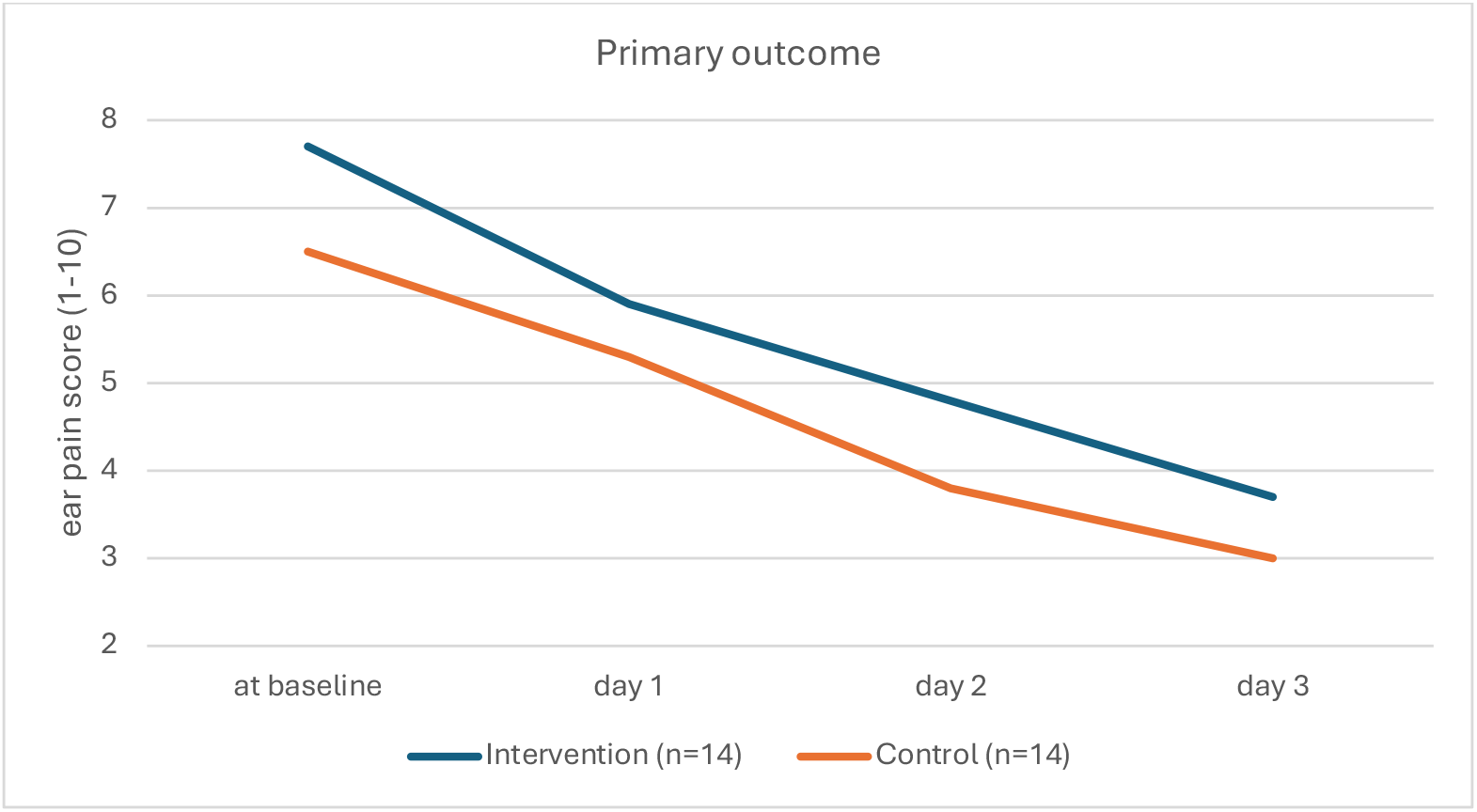
Mean ear pain scores over the first three days

### Secondary outcomes

Results for the secondary outcomes are summarized in Table 3. During the first 7 days after inclusion, 57.1% (8/14) children of the intervention group and 42.9% (6/14) of the control group were given antibiotics, for oral analgesics these figures were 78.6% and 100%, respectively. Mean total number of days with ear pain during the 4 weeks of follow-up was 6.5 in the intervention group and 4.6 days in the control group, GP re-consultations were 7 vs. 18, respectively.

**Table 3.**
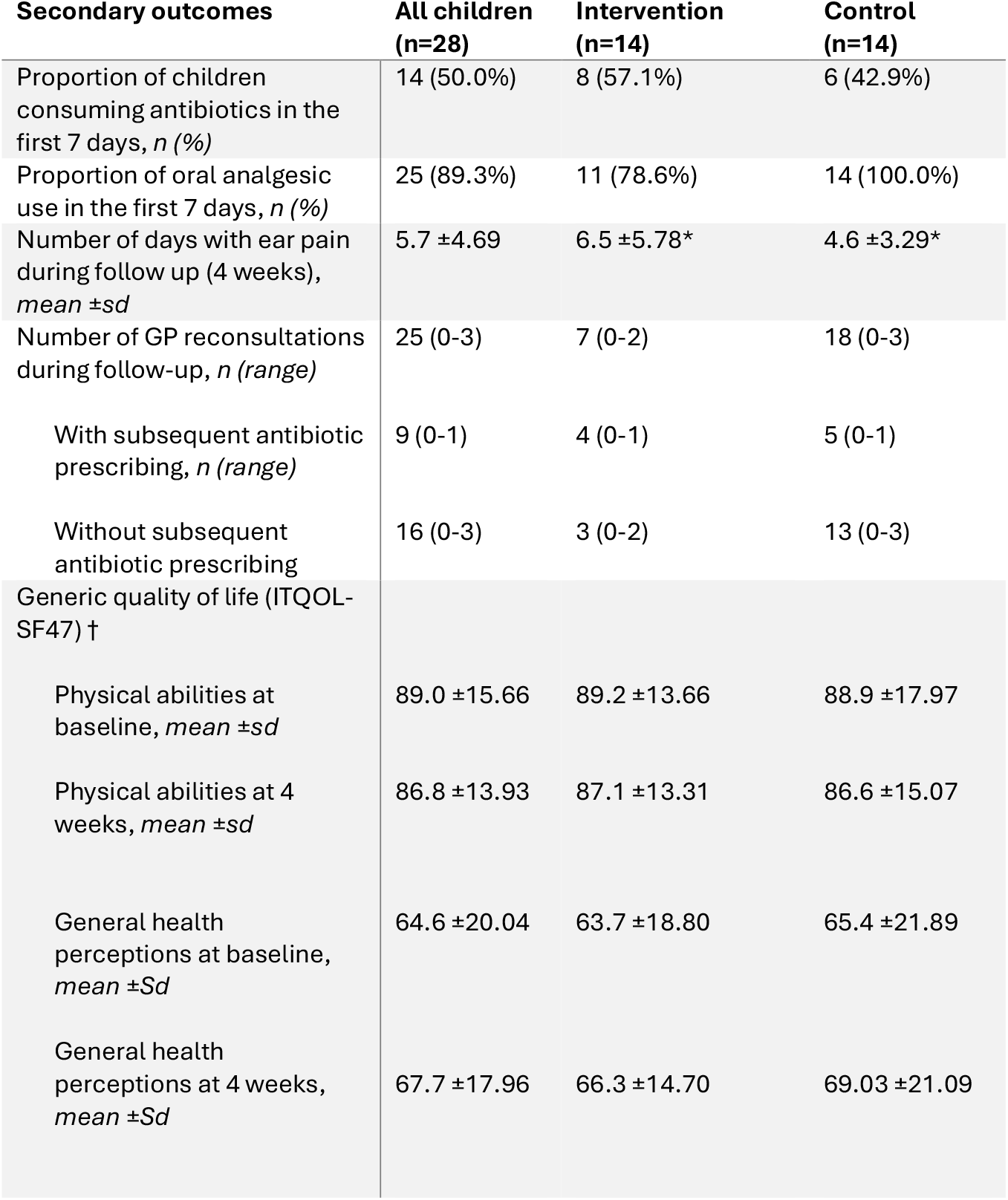
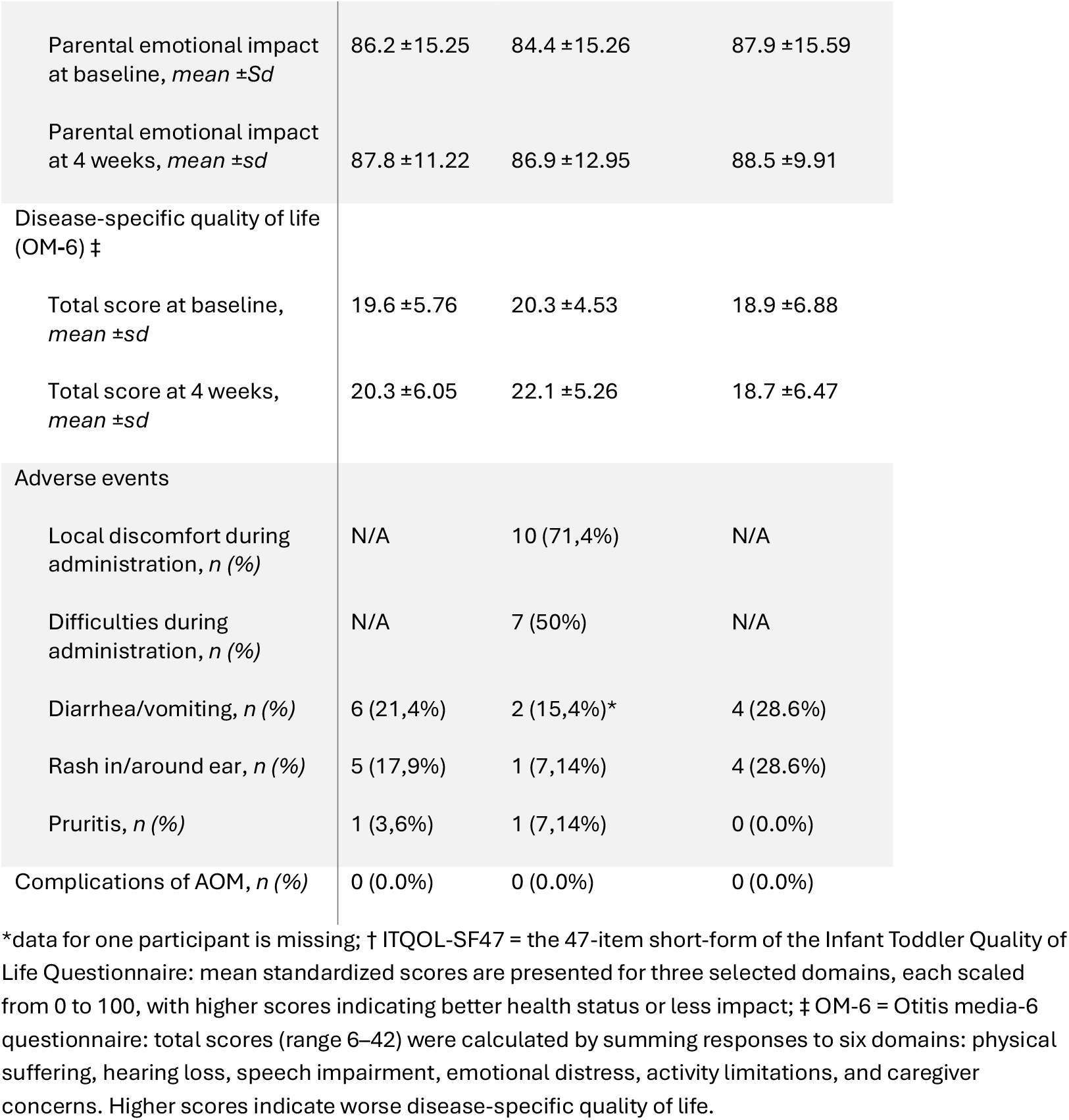
Secondary outcomes.

#### Overall symptom burden

The mean daily total symptom burden scores during the first 7 days after inclusion are presented in Figure 3. Similar to the primary outcome, scores decreased over time in both groups and were generally higher in the intervention group at individual time points.

**Figure 3.**
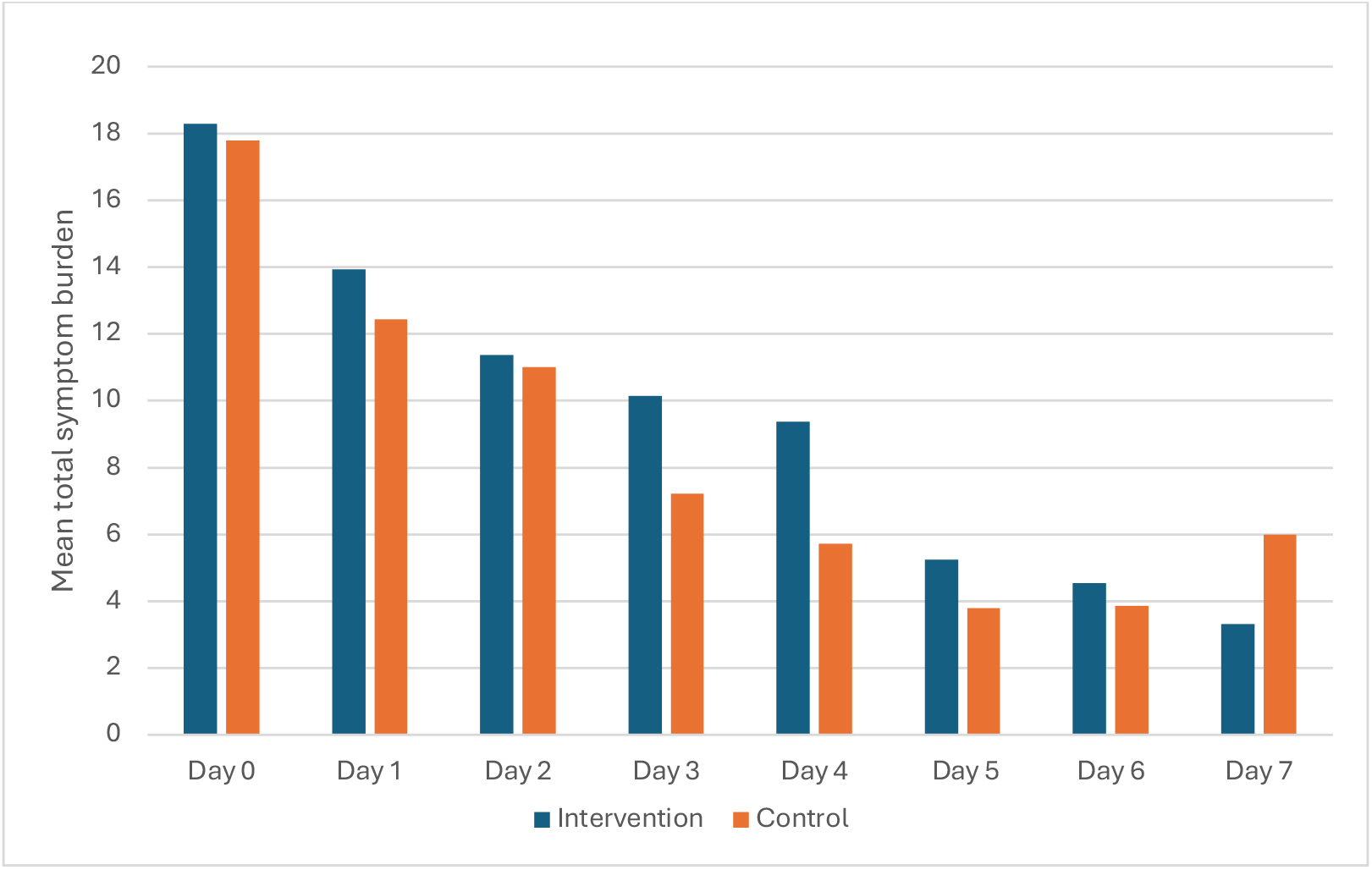
Mean total symptom burden in the first 7 days

#### Adverse events

One unrelated serious adverse event occurred in the intervention group, a participant was hospitalized on day 8 for acute appendicitis. Local discomfort and difficulty during administering the analgesic ear drops were reported in 71.4% (10/14) and 50% (7/14) of children in the intervention group, respectively. Other adverse events occurred at similar rates in the two groups (Table 3).

## DISCUSSION

### Main findings

Trial data were limited due to premature termination of the trial. No significant difference in mean parent-reported pain scores over the first three days was observed between children receiving analgesic ear drops in addition to usual care and those receiving usual care alone.

### Comparison with existing literature

To date, evidence supporting the use of analgesic ear drops in children with AOM is limited. A Cochrane review on this topic included five studies of which two (117 children) compared analgesic ear drops (phenazone/benzocaine and 2% lignocaine) to placebo. The authors concluded that these trials provide limited evidence that a single dose of analgesic ear drops is effective in reducing ear pain 30 minutes after administration in older children (aged 3 to 19 years) with AOM (18).

Since this review, two additional trials have been conducted. One UK-based trial compared repeated dosing of benzocaine 14 mg/ml and phenazone 54 mg/ml otic solution to placebo ear drops and usual care in children with AOM presenting in general practice who were not requiring immediate antibiotics. This trial was also ended prematurely due to operational challenges. The results of 84 participants (out of the planned 501) showed that fewer children in the analgesic ear drops group received antibiotics during the first 8 days after recruitment compared to those receiving usual care. A post hoc analysis further suggested that analgesic ear drops were associated with reduced ear pain on day 2 after recruitment compared to usual care.(14)

In a more recent trial based in Turkey 184 children with bilateral AOM were randomised into four groups: paracetamol plus 1% lidocaine ear drops, paracetamol alone, ibuprofen plus 1% lidocaine ear drops, or ibuprofen alone. Ten minutes after administration of ear drops, both lidocaine ear drop groups had significantly lower pain scores than their respective control groups.(16)

### Reflections

Recruitment proved the major challenge in this trial. Recruitment began during the COVID-19 pandemic, a period marked by a substantial decline in AOM incidence and a significant increase in GP workload due to ongoing pandemic demands.(25, 26) The high workload reduced the capacity of GPs to engage in research activities, while the decline in AOM incidence created an additional challenge by reducing the number of eligible patients available for inclusion. These phenomena have affected trials globally.(27) We experienced that recruitment remained a challenge after the COVID-19 restrictions were lifted. This likely reflected general barriers to enrolling participants in trials in general practice, such as lower-than-expected incidence rates, disruption of routine clinical workflow due to research activities, patient treatment preferences and perceived participant burden.(28, 29) A survey among investigators of 78 Dutch studies conducted in general practice found that only 46% reached their recruitment targets within the planned timeframe. This rate was 28% for trials focusing on incident cases, like our trial, which requires GPs to be alert to potential participants during their clinics and raise the opportunity to take part in the trial.(30)

We had included several strategies to address these challenges in our trial protocol, including minimising GP time with all trial procedures being conducted by the trial doctor, reimbursing GPs for patient identification and referral, co-designing recruitment processes and materials with a parent panel, engaging GP trainees, and regularly expanding and updating the network of participating general practices. These efforts proved insufficient to achieve the target sample size.

Other aspects of our trial design warrant reflection. Our choice to focus on self-reported outcomes was driven by clinical relevance, reflecting how symptoms are assessed and managed in routine care and thereby enhancing external validity. However, this choice as well the choice not to include placebo ear drop may have introduced outcome assessment bias. Nonetheless, this pragmatic design closely mirrors routine clinical practice, providing insight into the real-world effectiveness of analgesic ear drops.

### Implications for practice and future research

To provide more robust estimates of the effectiveness of analgesic ear drops for this common condition, further research is needed. We plan to combine the trial data with original data from other existing trials conducted so far in an individual participant data meta-analysis to strengthen the evidence base.

This trial highlights key challenges in conducting research on incident cases in primary care, particularly regarding recruitment. Understanding and addressing these barriers to develop collaborative and viable recruitment infrastructures for general practice research will be key to success of future trials in this setting.

## Data Availability

All data produced in the present study are available upon reasonable request to the authors

## DECLARATIONS

### Ethics approval

The trial protocol was approved by Medical Research Ethics Committee NedMec (21-447/G-D).

### Competing interests

The authors declare that they have no competing interests.

### Funding

The study is supported by a grant from the Netherlands Organisation for Health Research and Development (ZonMw– grant number 10060011910003). The funding agency had no role in the design, and will not have any role during execution of the trial, data analyses, interpretation of the data or decision to submit results.

### Data availability

All data produced in the present study are available upon reasonable request to the authors

## BOXES

### Box 1. Inclusion and exclusion criteria

#### Inclusion criteria

- Age 1 to 6 years
- Parent-reported ear pain in 24 hours prior to enrolment
- GP-diagnosis of (uni-or bilateral) AOM

#### Exclusion criteria

#### Children

- with (suspected) tympanic membrane perforation or ventilation tubes
- with ear wax obscuring visualisation of the tympanic membrane
- who are systemically very unwell or require hospital admission (e.g. child has signs and symptoms of serious illness and/or complications such as mastoiditis/meningitis)
- who are at high risk of serious complications including children with known immunodeficiency other than partial IgA or IgG2 deficiencies, craniofacial malformation including cleft palate, Down syndrome and previous ear surgery (with the exception of ventilation tubes in the past)
- who have a known allergy or sensitivity to study medication or similar substances (e.g. other amide-type anaesthetics: bupivacaine, mepivacaine, prilocaine, etc)
- who have taken part in any research involving medicines within the last 90 days, or any other AOM-related research within the last 30 days
- who suffer from chronic recurrent pain of another origin than the ear
- who have participated in this trial during prior AOM episode

## Notes

### Competing Interest Statement

The authors have declared no competing interest.

### Clinical Trial

The Netherlands Trial Register; NL9500. Date of registration: 28 May 2021.

### Clinical Protocols

https://pubmed.ncbi.nlm.nih.gov/36813504/

### Author Declarations

The trial protocol was approved by Medical Research Ethics Committee NedMec (21-447/G-D).

